# Stakeholder Networks and Systems for Health Equity: Accessibility of Newly Commissioned District Hospitals in Ghana

**DOI:** 10.1101/2025.01.31.25321475

**Authors:** Felix Frimpong, Charles Peprah, Ebenezer Owusu-Addo

**Author notes:** Corresponding Author: Corresponding author: Felix Frimpong.

## Abstract

Equitable access to healthcare necessitates the establishment of resilient stakeholder networks and comprehensive systemic integration to address the multifaceted needs of heterogeneous populations effectively. This research critically assesses the significance of stakeholder engagement in the planning and the provision of newly commissioned district hospitals in Ghana’s Ashanti Region, examining the extent to which their contributions influenced dimensions of accessibility and the uptake of services.

A convergent parallel mixed-method design was employed, incorporating qualitative methods (indepth interviews) with stakeholders such as healthcare providers, community leaders, and local officials, and quantitative data was collected from healthcare seekers. The study applied an accessibility framework—availability, affordability, accommodation, geographical access, and acceptability—to assess the impact of systemic and stakeholder-driven initiatives.

Stakeholder networks played a crucial role in the strategic planning and provision of the newly commissioned district hospital services. The collaborative decision-making process facilitated the key dimensions of access to healthcare to be prioritized. Nevertheless, the issue of affordability surfaced as a systemic limitation that critically impacted service utilization, particularly among economically disadvantaged populations. The research findings indicated that demographic groups characterized by female gender, advanced age, and higher educational attainment derived greater benefits from the stakeholder-oriented planning methodologies, whereas rural populations and those belonging to socio-economically disadvantaged backgrounds encountered substantial obstacles.

The findings underscore the critical role of stakeholder networks in health promotion. Their engagement ensured alignment between systemic goals and community needs, fostering service uptake and reducing disparities. The study emphasizes the importance of leveraging multi-level connections between policy, health systems, and community networks to achieve Universal Health Coverage and Sustainable Development Goal 3

## Background of study

The provisions of social infrastructural facilities have been crucial for the growth of the economy and society in recent years (Pascale et al. 2020). They are mostly used to solve infrastructure capacity issues or to open new economic opportunities (Kumaraswamy et al. 2017; Ninan et al. 2020). In addition to civil infrastructure, another category of urban infrastructure known as social infrastructure exists (Dyer et al. 2020). This form of infrastructure is essential for the advancement of society and facilitates the establishment of services such as healthcare facilities, among other things, to promote population health (Ninan et al. 2020). However, most of the implemented methods often fail to meet the requirements of end-users. The outcome of a study established that over 70% of organizations start projects that do not meet the expectations of stakeholders or fail to yield the needed profit (Prebanic and Vukomanović, 2021; Nguyen et al. 2021).

Studies that look at the type of stakeholder engagement in intricate building and infrastructure projects often adopt two major methods. The first strategy specifically addresses the stakeholder management procedure as carried out by another study (Bahadorestani et al., 2020). The second category of research examines stakeholder engagement via the lenses of institutional, organizational, and complexity theory (Kumaraswamy et al. 2017; Rezvani et al. 2018; Ninan et al. 2020; Wagner et al. 2022). There is widespread agreement that in major infrastructure projects, stakeholder involvement is crucial (Cuppen et al., 2016; Pascale et al., 2020; Prebanic and Vukomanović, 2021; Nguyen et al., 2021).

Therefore, to meet the requirements of end-users, most government institutions often involve stakeholders when implementing a project from the initiation and planning phase through the implementation stage (see Dyer et al. 2020). The reverse is what happens during the implementation of major projects in developing countries, especially in Sub-Saharan Africa. Stakeholder management theory states that projects are effective when they consider stakeholders’ needs and requirements through the stakeholder management process (Pascale et al. 2020).

In Ghana, the uneven distribution of hospital accessibility, in addition to limited district hospitals and other fiscal constraints, calls for better planning for the next generation in terms of accessibility to healthcare facilities (Falchetta et al. 2020). Many countries, including Ghana, have developed and introduced healthcare services and policies (Sustainable Development Goals (SDGs) target 3.8) by 2030) to improve access to healthcare (Kwarteng et al. 2020).

One of the tenets of the policy requires the provision of new healthcare facilities to increase access to healthcare across Ghana (Asare-Akuffo et al. 2020). Therefore, the government of Ghana established district hospital projects across Ghana (Haruna et al. 2019). One of the beneficiary regions in Ghana is the Ashanti Region. In the Ashanti region, some health facilities are built 40 km away from communities (Banke-Thomas et al. 2019). As a result, accessibility to these health facilities and services is affected by socioeconomic factors, including long distances to health facilities, poor infrastructure delivery planning, and inadequate community involvement and empowerment (Buor, 2003; Asare-Akuffo et al. 2020).

The aforementioned factors, among other factors, affect most of the population, who depend on district hospitals as the first referral point for their healthcare needs and access their right to healthcare services. This results in some district hospitals being available but not accessible (Howard and Rhule, 2021). According to Ghana’s Universal Health Coverage Policy of 2020, the provisions of any healthcare facility need to be accompanied by stakeholder engagement from the project planning stage to the implementation stage. However, a preliminary investigation revealed that there was little to no stakeholder engagement during the establishment of the newly commissioned district hospitals in the Ashanti Region. This study sought to assess the extent of stakeholder engagement during the provisions of the newly commissioned district hospitals in the Ashanti Region of Ghana.

## 2.0 METHODS AND MATERIALS

### 2.1 Scope of the Study

The study was conducted in three districts of the Ashanti Region in Ghana where there has been a new provision of district hospitals to increase the number inequality of populace access to healthcare in the country (see Figure 2.1). The selected districts are Ahafo Ano North Municipality, Bekwai Municipal Municipality, and Asante Akim Central Municipality. The Ahafo Ano North Municipality is located in the Ashanti Region of Ghana. Ahafo Ano North municipality is among the 43 administrative districts in the Ashanti Region. Municipal status was granted to it in November 2017 (L.I. 2264), after which it had previously been a district (Municipal Coordinating Unit, Ahafo-Ano North Municipal Assembly, 2022). Ahafo-Ano North is bordered by the Tano North and South, Asutifi South, Ahafo Ano South East, and Ahafo Ano South West districts (Municipal Coordinating Unit, Ahafo-Ano North Municipal Assembly, 2022).

**Figure 2.1:**
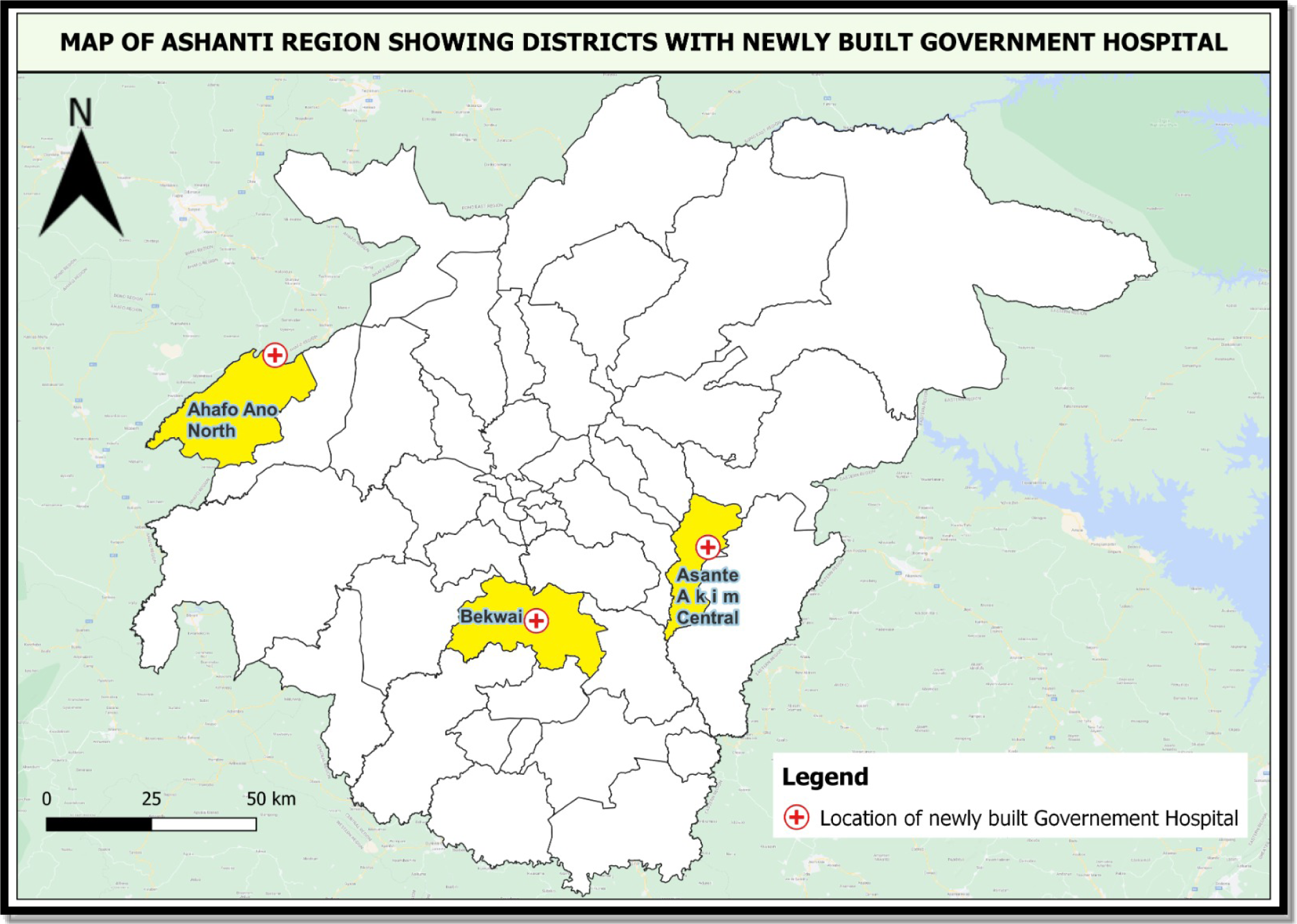
Selected study districts. Source: Author’s Construct, 2024.

### 2.2 Research Approach

A quantitative approach was adopted for the study. Given Creswell (2009), quantitative research is the inquiry into a social problem, which is based on testing a hypothesis or theory composed of variables, measured with numbers, and analyzed with a statistical procedure. Quantitatively, the study assessed stakeholders’ perceptions, attitudes, and experiences of their involvement in the planning process of newly established district hospitals in the Ashanti Region of Ghana. Therefore, primary data was collected through a survey by administering structured questionnaires. Generally, the attitudes, perceptions, and experiences of stakeholders based on their involvement in the hospital planning process were assessed via a five-point Likert scale where 1= Strongly Disagree, 2= Disagree, 3 = Uncertain, 4 = Agree, and 5 = Strongly Agree.

### 2.3 Data needs and sources

The study depended on both secondary information and primary sources of data. Secondary information on the importance of stakeholder engagement in project implementation, challenges of stakeholder engagement in project planning and implementation, and types of stakeholders and their influence and power on project planning and implementation were retrieved from the available relevant literature, such as articles, journals, organizational reports, and institutional websites. On the other hand, primary data was gathered through a survey. The research instrument used was a close-ended questionnaire. The types of primary data gathered include the perceptions, attitudes, and experiences of stakeholder engagement in the provision of newly commissioned district hospitals in the Ashanti Region of Ghana.

### 2.4 Sampling Technique

The study population included key stakeholders who were consulted during the planning stage of the district hospital project. The key stakeholders consulted are Hospital Administrators, Traditional Authorities, Assembly Members (elected and appointed members), Estate Officers, and District Spatial Technical Sub-Committee Members (Physical Planning Department, District Development Planning Officer, Works Department, Roads Unit of the District Assembly, Disaster Prevention Department, Lands Commission, Environmental Protection Agency, District Fire Officer, District Health Department, Two Chairpersons of the Zonal Council, Electricity Company of Ghana). The sampling frame of the study is 356; Bekwai Municipality=154; Ahafo Ano North District=113; Asante Akim Central Municipality=89. Based on the sampling frames, sample sizes were determined via the mathematical model of Miller and Brewer (2003), where n is the sample size, N is the sample frame, and α is the margin of error. A confidence level of 95% with a margin error of 5% was allowed for the study to reflect the true representation of the phenomenon studied.

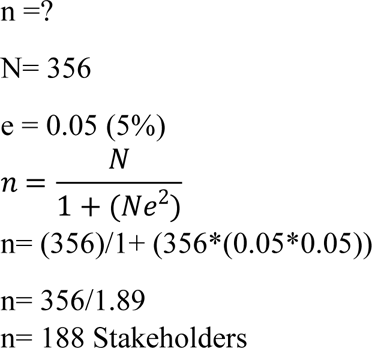

Using sample proportions, the number of stakeholders selected from each district was estimated to be 81 from Bekwai Municipality, Ahafo Ano North District (60), and Asante Akim Central Municipality (47).

### 2.5 Sampling Technique

A simple random sampling technique was used to select key stakeholders for the study. Using a simple random sampling technique, each member of the survey population possessed an equal chance of being selected to be part of the sample for the study. A simple random technique is mostly used to eliminate or reduce the possible occurrence of biases from the selection process, which normally results in the representativeness of the sampled population, leading to the generalization of the research findings (Rahman, 2023).

To avoid selection bias when the simple random sampling technique is used, a random number generator (RNG) was employed to facilitate the selection process to allow for equal and nonzero chances of each house being selected (Bhattacharjee & Das, 2019). Generally, a probability sampling method is used in selecting a sample based on random selection procedures in which every member of the target population has a known, non-zero chance of being included in the sample (Rahman, 2023).

### 2.6 Data collection methods and instruments

The structured interview method was used for data collection. Therefore, a close-ended questionnaire was used for data collection. Close-ended questionnaires contain a list of options from which the respondent is expected to choose (Sreejesh et al. 2014). In this study, the questionnaires were arranged in four sections. The first section covered the demographics of the respondents, the second section covered the perceptions of stakeholders regarding their involvement in the planning of district hospitals, and the third section covered the attitudes of stakeholders regarding their involvement in the planning of district hospitals. The final and fourth sections covered the experiences of the influence of stakeholders on the planning involvement of district hospitals.

### 2.7 Data processing and analysis

After the administration and retrieval of the administered questionnaires, the data were cleaned, and SPSS version 20 software was used for the final analysis. The data gathered were coded numerically to make the analysis more manageable and accurate. The data were analyzed via descriptive statistics (means, standard deviations, frequencies, percentages).

The data on the demographic characteristics of the respondents were analyzed via frequency and percentages and are presented in tables. For the attitudes, perceptions, and experiences of stakeholders, the data were analyzed using means and standard deviations. Primary data collected via the Likert scale are best analyzed via means and standard deviations (see Sullivan & Artino, 2023).

### 2.8 Ethical Considerations

These ethical considerations of the study included informed, voluntary consent, confidentiality of information, and respondent anonymity, aligning with the General Research Ethics Board guidelines of the Kwame Nkrumah University of Science and Technology (KNUST). Ethical approval was obtained from the Kwame Nkrumah University of Science and Technology (Humanities and Social Research Ethics Committee) **Ref. No: HUSSREC/AP/30/Vol.3**.

After the Department of Planning at KNUST approved the ethics application for conducting surveys, interviews, participant observations, and secondary data collection, the research commenced with an explanation of its purpose to involved agencies, institutions, and community study sites. Following departmental requirements, semi-structured interview schedules, questionnaires, and guides were prepared, including introductory letters outlining the study’s goals and ensuring participant confidentiality and anonymity.

Ethical consideration was very important in this research because it promoted an environment of trust, accountability, and mutual respect among the researcher and the respondents. For example, the respondents did not permit the taking of photographs during the discussion. Finally, the collected data were not falsified to paint a different picture of the issues being studied.

## 3.0 RESULTS

### 3.1 Demographic information of the respondents

The demographic characteristics of the respondents are presented in Table 3.1. The study revealed that the majority (94%) were male, and the remaining 6% were female. Additionally, most of the respondents (39.0%) indicated that they had attained a master’s degree, 33.3% indicated that they had attained a diploma, and the remaining 28.0% indicated that they had attained a degree. Finally, most of the respondents (43.3%) reported that they are government employees; 30.7% of the respondents indicated that they are traders, with the remaining 26.0% being artisans, such as carpenters, masons, tailors, and hairdressers.

**Table 3.1:** Demographic characteristics.

| Demographics | Frequency(n) | Percentage (%) |
| --- | --- | --- |
| **Sex** |  |  |
| Male | 335 | 94.0 |
| Female | 21 | 6.0 |
| **Age (years)** |  |  |
| 25-29 | 164 | 46.0 |
| 30-45 | 167 | 47.0 |
| 46-60 | 25 | 7.0 |
| **Highest Education** |  |  |
| Diploma | 117 | 33.0 |
| Degree | 100 | 28.0 |
| Masters | 139 | 39.0 |
| **Job Designation** |  |  |
| Trading | 110 | 31.0 |
| Artisan | 93 | 26.0 |
| Employed by government | 153 | 43.0 |

**Source: Field Survey, 2024**

### 3.2 Types of stakeholders involved in the planning of the hospital project

Different categories of stakeholders were involved in planning the newly commissioned district hospitals in the Ashanti Region. The types of stakeholders engaged in the hospital planning process include Hospital Administrators, Traditional Authorities, Assembly Members (elected and appointed members), Estate Officers, and District Spatial Technical Subcommittee Members (Physical Planning Department, District Development Planning Officer, Works Department, Roads Unit of the District Assembly, Disaster Prevention Department, Lands Commission, Environmental Protection Agency, District Fire Officer, District Health Department, Two Chairpersons of the Zonal Council, Electricity Company of Ghana) (see Figure 3.1).

**Figure 3.1:**
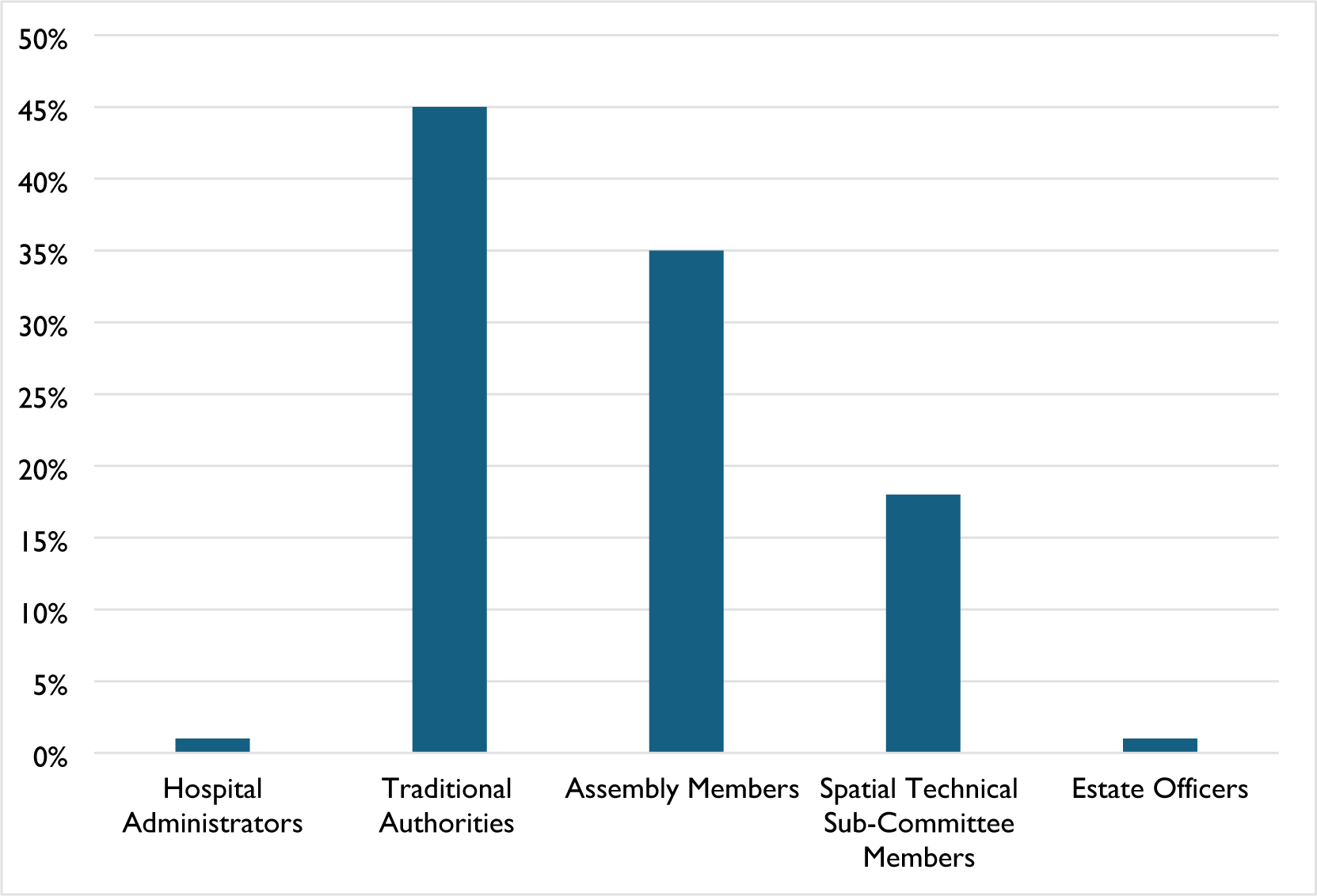
Categories of stakeholders engaged. Sources: Field Survey, 2024.

### 3.3 Stakeholders’ perception of involvement in the hospital planning process

Stakeholders’ perceptions of their involvement in the hospital planning process were assessed via a five-point Likert scale where 1= Strongly Disagree, 2= Disagree, 3 = Uncertain, 4 = Agree, and 5 = Strongly Agree.

Table 3.2 presents the descriptive statistics of stakeholders’ perceptions of the hospital planning process. Most (mean=4.62, SD=0.30) of the respondents held the view that they had adequate opportunity to participate in the hospital planning process, followed by those who (mean=4.54, SD=0.50) believed that they received enough information about the progress of hospital planning, followed by most (mean=4.52, SD=0.88) of the respondents, who believed that their opinions and concerns were considered in the hospital planning process. Additionally, the majority (mean=4.50, SD=0.61) believed that the hospital planning process was transparent, whereas a minority (mean=4.24, SD=1.10) believed that stakeholder engagement in the hospital planning process was significant for successful completion. In general, the respondents had good perceptions of the hospital planning process.

**Table 3.2:**
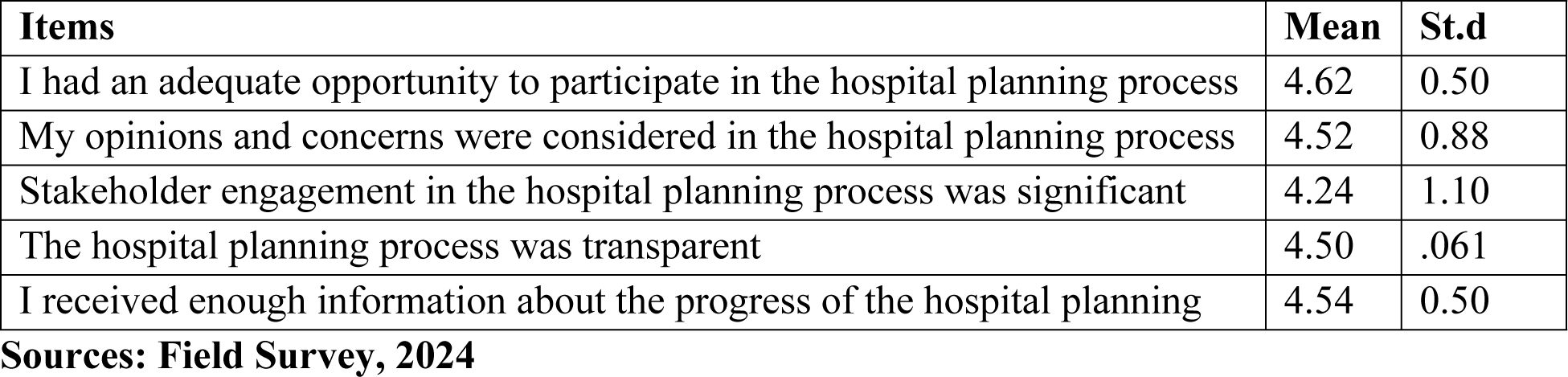
Stakeholders’ perceptions of the hospital planning process.

### 3.4 Stakeholders’ attitudes toward involvement in the hospital planning process

Stakeholders’ attitudes toward their involvement in the hospital planning process were assessed via a five-point Likert scale where 1= Strongly Disagree, 2= Disagree, 3 = Uncertain, 4 = Agree, 5 = Strongly Agree. As shown in Table 3.3 below, most (mean=4.62, SD=0.49) of the respondents stated that their willingness to support the full planning of the hospital followed by planning for the new hospital is essential for healthcare users (mean=4.57, SD=0.49), followed by most (mean=4.54, SD=0.53) of the respondents, who stated that they actively advocated for the hospital planning process (mean=4.54, SD=0.53). Others also expressed their willingness to contribute my resources to support hospital planning (mean=4.47, SD=0.94), with a minority (mean=4.34, SD=1.24) being concerned with the successful planning of the new hospitals. In general, the respondents had a positive attitude towards the hospital planning process.

**Table 3.3:** Stakeholders’ attitudes towards the hospital planning process.

| Items | Mean | St.d |
| --- | --- | --- |
| Planning for the new hospital was essential for healthcare users | 4.57 | 0.49 |
| I actively advocated for the hospital planning process | 4.54 | 0.53 |
| I was willing to support the full planning of the hospital | 4.62 | 0.49 |
| I was willing to contribute my resources to support hospital planning | 4.47 | 0.94 |
| I was concerned about the successful planning of the hospital | 4.34 | 1.24 |

**Sources: Field Survey, 2024**

### 3.5 Experiences of the stakeholders involved in the hospital planning process

The experiences of stakeholders involved in the hospital planning process were assessed via a five-point Likert scale where 1= Strongly Disagree, 2= Disagree, 3 = Uncertain, 4 = Agree, and 5 = Strongly Agree. Table 3.4 presents descriptive statistics on the experiences of the stakeholders involved in the hospital planning process. As shown in the table below, most (mean=4.60, SD=0.49) of the respondents stated that they enjoyed being part of the hospital planning process, followed by making valuable contributions to hospital planning (mean=4.57, SD=0.48). Additionally, the majority (mean=4.56, SD=0.51) of the respondents stated that being part of the hospital planning process broadened their understanding of the project planning process, followed by being actively involved in hospital planning (mean=4.53, SD=0.55), with the lowest number (mean=4.48, SD=0.94) of respondents stating that their views were taken into consideration. Overall, the respondents had good experience engaging in the hospital planning process (mean=4.60, SD=0.55).

**Table 3.4:** Experiences of stakeholders involved in the hospital planning process.

| **Items** | **Mean** | **St.d** |
| --- | --- | --- |
| I enjoyed being part of the hospital planning process | 4.60 | 0.49 |
| I made valuable contributions to the hospital's planning | 4.57 | 0.48 |
| I was actively involved in the hospital planning | 4.53 | 0.55 |
| My views were taken into consideration | 4.48 | 0.94 |
| Being part of the hospital planning process broadened my understanding | 4.56 | 0.51 |

**Sources: Field Survey, 2024**

## 4.0 DISCUSSIONS

### 4.1 Types of stakeholders involved in the planning of the hospital project

In general, the various stakeholders involved came from diverse backgrounds with rich experience that was essentially relevant to the successful planning of healthcare facilities. The diversity of the stakeholders encouraged the sharing of ideas relevant to the successful planning of healthcare facilities. The diverse backgrounds of the participants also highlight their influence and power over the project. Among all the stakeholders involved, only traditional authorities had much power and influence over the project implementation process. The establishment of district hospitals in the Ashanti Region is a national-level policy; hence, local-level stakeholders (except traditional authorities) have little say in overall project implementation. However, the power and influence of traditional authorities come into play when deciding the location for the eventual construction of health facilities to ensure easy physical accessibility for healthcare seekers. In Ghana, traditional authorities are custodians of approximately 80% of land and hence influence key land use planning functions in the form of land use determination and management (see Siiba et al. 2018; Boamah & Amoako, 2020). By extension, traditional authorities have the power and influence to determine the easy physical accessibility of newly commissioned district hospitals. Therefore, the involvement of traditional authorities at the planning stage of the project is essential for ensuring that health facilities are situated at the optimum location for healthcare seekers.

### 4.2 Stakeholders’ perceptions of their involvement in the hospital planning process

In general, the respondents had good perceptions of the hospital planning process. Therefore, their engagement expedited the hospital planning process. Thus, a positive perception of stakeholders about a project is essential for its successful completion. In this case, perception refers to the procedure of achieving awareness or comprehension of sensory information (see Bai, 2001). This refers to the mental interpretation of physical sensations created by stimuli from the outside world; mental interpretation in this context refers to the process of creating an internal model of the environment (Li and Wu, 2005).

The good perception of stakeholders concerning their involvement in the hospital planning process was due to their active involvement in the entire process, leading to the participatory planning process. Thus, a participatory project planning process guarantees local ownership of a project and hence the commitment to ensure the success of the project, as is the case for newly commissioned district hospitals in the Ashanti Region (Khan, 2021). This underscores the notion that stakeholder involvement in the initiation and planning process of a project is significant. According to Paddock (2013), the advantages associated with participatory project planning include the creation of a mechanism for receiving feedback and ideas for counteractive actions, making the project adaptable, and strengthening local ownership. When the project planning process is participatory, it redistributes power for decision-making purposes and gives this power to individuals who are direct beneficiaries of the project (see Mulwa, 2008). The participatory project planning process acknowledges that local people have knowledge and experience with the success of the project. This was the case during the planning stage of the newly commissioned district hospitals in the Ashanti Region.

### 4.3 Stakeholders’ attitudes toward involvement in the hospital planning process

Generally, the respondents had a positive attitude towards the hospital planning process. It follows that the stakeholders were willing to be flexible to ensure that the project was planned successfully. Stakeholders’ attitudes towards this project ranged from actively supporting it with all their resources to willingly engaging with other stakeholders to ensure the effective planning phase of the project. This is because the stakeholders viewed the hospital project as an important social infrastructure that is required for their well-being. The implication is that the attitudes of the stakeholders shaped their thoughts, emotions, and worldviews towards the success of the projects. Research indicates that a venture’s success or failure affects people’s perceptions of it (see Aas et al. 2005). Therefore, a person is more likely to exhibit positive attitudes toward the accomplishment of a purpose if she is more open-minded toward it (see Frederick, 2005). Therefore, it can be argued that the open-mindedness of the stakeholders resulted in their readiness to support a worthwhile endeavor (district hospital project) (see Aas et al. 2005; Frederick, 2005).

### 4.4 Experiences of stakeholders involved in the hospital planning process

Overall, the respondents had good experience engaging in the hospital planning process during the establishment of the newly commissioned hospitals in the Ashanti Region. This is because the stakeholders had good working relationships when planning the hospitals. Good working relationships among stakeholders during a project lead to good working experience (Aaltonen and Kujala, 2016). Numerous stakeholders are connected in the social context of the project’s implementation when they engage with each other (see Turner and Müller, 2017). In addition to good working relationships, mutual trust among stakeholders is another contributing factor to the good experience of stakeholders. Owing to the level of trust, each of the respondents shared their thoughts and opinions regarding the successful planning of the district hospitals. Trust is key among stakeholders during the implementation of a project (Turner and Müller, 2017). The implication is that good relationships and trust are key to ensuring a positive experience during the implementation of a project.

## 5.0 CONCLUSION

The study assessed stakeholder engagement in the provision of newly commissioned district hospitals. The objective of the study includes examining the types of stakeholders engaged as well as assessing the perceptions, attitudes, and experiences of stakeholder engagement in the initiation and planning of a newly commissioned district hospital. The study established different categories of stakeholders involved in the planning of the newly commissioned district hospitals in the Ashanti Region to evoke diversity in the sharing of ideas at the initiation and planning stages of the district hospitals. In general, the respondents had good perceptions of the hospital planning process. Additionally, the respondents had a positive attitude towards the hospital planning process. Finally, overall, the respondents had good experience engaging in the planning process of the new district hospitals. The study established that creating a good working environment during the implementation of a project is essential for influencing the perceptions, attitudes, and experiences of key stakeholders engaged in the project planning stage. Additionally, local-level stakeholders that wield much power and influence need to be prioritized when they are engaged during the implementation of state-led projects. Finally, the engagement of stakeholders in a project should be prioritized as a policy requirement when major infrastructural projects are implemented. The engagement of relevant local-level stakeholders ensures that the implemented projects meet the requirements of the end-users. Stakeholders’ involvement in the implementation of district hospitals is key to achieving the aims and objectives of the Universal Health Coverage Policy of 2020. The results indicate that heightened stakeholder engagement from the initiation phase of projects could significantly enhance healthcare access and equity in developing nations such as Ghana, which is key to achieving the aims and objectives of Sustainable Development Goal Three.

## Declaration

### Ethics approval and consent to participate

Ethical approval was obtained from the Kwame Nkrumah University of Science and Technology (Humanities and Social Research Ethics Committee) **Ref. No: HUSSREC/AP/30/Vol.3.**

### Consent for publication

Not Applicable

### Availability of data and materials

Data for the study are available and can be released following a reasonable request by writing to the corresponding author.

### Authors’ contributions

Conceptualization, F.F., C.P., and E.O-A.; Data collection, F.F., an C.P.; Data Analysis, F.F.; Writing – original draft, F.F., C.P., and E.O-A.; Methods, F.F., C.P., and E.O-A.; Writing – review and editing, F.F., C.P., and E.O-A. All authors have agreed to the published version of the manuscript.

## Data Availability

Data for the study is available upon request

## Acknowledgment

We would like to acknowledge the staff of the three district hospitals for their support in completing this research. Our appreciation also goes to the enumerators who supported the data collection.

